# Strategic vaccine stockpiles for regional epidemics of emerging viruses: a geospatial modeling framework

**DOI:** 10.1101/2024.01.19.24301505

**Authors:** Colin J. Carlson, Romain Garnier, Andrew Tiu, Stephen Luby, Shweta Bansal

## Abstract

Multinational epidemics of emerging infectious diseases are increasingly common, due to anthropogenic pressure on ecosystems and the growing connectivity of human populations. Early and efficient vaccination can contain outbreaks and prevent mass mortality, but optimal vaccine stockpiling strategies are dependent on pathogen characteristics, reservoir ecology, and epidemic dynamics. Here, we model major regional outbreaks of Nipah virus and Middle East respiratory syndrome, and use these to develop a generalized framework for estimating vaccine stockpile needs based on spillover geography, spatially-heterogeneous healthcare capacity and spatially-distributed human mobility networks. Because outbreak sizes were highly skewed, we found that most outbreaks were readily contained (median stockpile estimate for MERS-CoV: 2,089 doses; Nipah: 1,882 doses), but the maximum estimated stockpile need in a highly unlikely large outbreak scenario was 2-3 orders of magnitude higher (MERS-CoV: ∼87,000 doses; Nipah ∼1.1 million doses). Sensitivity analysis revealed that stockpile needs were more dependent on basic epidemiological parameters (i.e., death and recovery rate) and healthcare availability than any uncertainty related to vaccine efficacy or deployment strategy. Our results highlight the value of descriptive epidemiology for real-world modeling applications, and suggest that stockpile allocation should consider ecological, epidemiological, and social dimensions of risk.

## Introduction

Recent decades have witnessed a marked increase in animal-to-human spillover of pathogens with epidemic and pandemic potential, likely as a result of changes in ecosystem function, agriculture, climate, and human-wildlife contact (Baker et al. 2022; Jones et al. 2008; Mahon et al. 2022; Carlson, Albery, et al. 2022; Keesing et al. 2010). Due to rising global mobility, a greater proportion of these outbreaks have become serious epidemics (Balcan et al. 2009; Loh et al. 2015; Redding et al. 2019; Marani et al. 2021; Smith et al. 2014). The 2012 emergence of the Middle East respiratory syndrome coronavirus (MERS-CoV) marked the start of a decade of prolonged multinational outbreaks of viruses that have conventionally been seen as readily contained by a rapid response (Fraser et al. 2004; Carlson, Boyce, et al. 2022). In the time since, this pattern has recurred in regional epidemics of Ebola virus (Hampton et al. 2023), explosive epidemics of mosquito-borne diseases in the Americas (Musso et al. 2018), the Covid-19 pandemic, and most recently, the 2022 global mpox outbreak (Alakunle and Okeke 2022).

The scale of these outbreaks—and the challenges of non-pharmaceutical interventions at those scales—have increasingly made vaccination the first (and sometimes only) line of defense. After the 2014 epidemic in West Africa, vaccines have become a standard component of Ebola virus outbreak response (Potluri et al. 2022; Wells et al. 2019; Worden et al. 2019); similarly, *Vaccinia*-based vaccines developed for smallpox are also effective against mpox (Poland, Kennedy, and Tosh 2022), and were instrumental in containing the 2022 outbreak. For novel pathogens, lineages, or variants, the speed of the R&D pipeline has been a subject of historical concern (World Health Organization 2017), but the Covid-19 pandemic has demonstrated the feasibility of comparatively rapid manufacturing and rollout, as well as the value of investments in novel vaccine platform technologies (Więcek 2022; Cleve 2021; Moghadas et al., n.d.). Anticipated progress towards the development of universal vaccines will further improve preparedness for novel pathogens (Arevalo et al. 2022; Cohen et al. 2021).

Proactive investments in vaccine stockpiles could help contain outbreaks earlier, and prevent larger epidemics and pandemics altogether. Equally important, they could reduce the drastic inequity that has resulted from Global North monopolization of intellectual property, manufacturing capacity, and vaccine allocation that has been a hallmark of both the Covid-19 and mpox responses. Though the optimal design of vaccine stockpiles is a familiar problem in quantitative epidemiology (Thompson and Duintjer Tebbens 2016; Tebbens et al. 2010; Matrajt, Halloran, and Longini 2013), less work has explored application to emerging pathogens (Lerch et al. 2022).

Here, we develop a stochastic geospatial modeling framework that accounts for multiscale dynamics of disease emergence, including spillover risk, spatial heterogeneity in population distribution and healthcare infrastructure, and spatial connectivity via human mobility. Focusing on two viruses with regional epidemic potential—Nipah virus and Middle East respiratory syndrome coronavirus (MERS-CoV)—we demonstrate how vaccine stockpiling can be approached as a spatial design problem, and highlight where future work can reduce model uncertainty.

## Methods

Our project was conducted in 2019 as a quantitative exercise in support of the Coalition for Epidemic Preparedness Innovations (CEPI), which coordinates a global effort to develop novel vaccines and vaccine technologies that can reduce epidemic and pandemic risk. Since 2017, CEPI has funded the development of several dozen vaccine candidates, with a core focus on six priority pathogens (Lassa virus, MERS-CoV, Rift Valley fever, Nipah virus, chikungunya virus, and Ebola virus), as well as (since 2020) Covid-19 and other coronaviruses yet to emerge. In service of that effort, our project simulated the vaccine stockpile needs that would arise in a major regional multi-national epidemic of two of these pathogens (Nipah virus and MERS-CoV). Our models are intended to reproduce the characteristics of real, historical outbreaks of these pathogens. While concerns have been raised that new lineages of either pathogen could arise with greater capacity for human-to-human transmission (Schroeder et al. 2021; Luby 2013), (Schroeder et al. 2021; Luby 2013), rather than high transmissibility scenarios in which global mass vaccination would be necessary. To address other aspects of uncertainty, we conduct sensitivity analysis on over a dozen epidemiological parameters, and highlight the impact of data gaps.

Features our focal pathogens share include (1) frequent zoonotic (animal-to-human) spillover, (2) an anticipated risk of unexpectedly-large regional outbreaks, and (3) ongoing vaccine development efforts. Nipah virus is a bat-origin virus that was first described from a 1998 outbreak in Malaysia; most reported outbreaks since have been in Bangladesh or occasionally India. Secondary transmission is limited but nontrivial, with outbreaks usually limited to a few dozen cases (Nikolay et al. 2019). Similarly, MERS-CoV also likely originated in bats (Anthony et al. 2017), but is maintained in domesticated camels. Though its capacity for human-to-human spread has so far been middling (particularly compared to SARS-CoV-2) (Dudas et al. 2018), outbreaks have reached over two dozen countries. Despite limited evidence of community spread, both pathogens have a notable track record of spreading in healthcare settings: in the 2015 outbreak of MERS-CoV, a single infected patient seeking treatment at multiple facilities in South Korea sparked an outbreak of 186 cases, including 25 healthcare workers (Lee et al. 2017; Hui et al. 2018); during the 2018 Kerala outbreak of Nipah virus, nosocomial transmission accounted for most or all secondary cases (Arunkumar et al. 2019), and more limited instances have been reported in smaller outbreaks (Tan and Tan 2001; Sazzad et al. 2013).

We focus our attention on a geographic region for each pathogen: for Nipah, our focal region consists of India and Bangladesh in the Indian subcontinent; for MERS-CoV, our focal region consists of 21 countries of the Middle East and Northeast Africa (Bahrain, Djibouti, Egypt, Eritrea, Ethiopia, Iran, Iraq, Israel, Jordan, Kuwait, Lebanon, Oman, Palestine, Qatar, Saudi Arabia, Somalia, South Sudan, Sudan, Syria, United Arab Emirates, and Yemen).

To estimate vaccine stockpile needs for these two pathogens, we developed a multilayer modeling framework that predicts local and regional epidemiological dynamics post-emergence. Our approach accounts for geographic heterogeneity in both spillover risk and human-to-human transmission, drawing on previous frameworks developed for Ebola virus and influenza (Redding et al. 2019; Pigott et al. 2017; Berger et al. 2018). As a novel extension, we added a third layer for vaccination, with multiple strategies represented. All simulations and analyses were conducted in Python 3.7 and R 3.5.

### Stage 1. Ecological risk factors for emergence

Fine-scale mapping of spillover risk poses a substantial challenge, particularly for pathogens with only a few dozen recorded animal-to-human transmission events. Machine learning is often used to predict hotspots of spillover intensity, including for Nipah (Deka and Morshed 2018; Peterson 2015; Walsh 2015) and MERS-CoV (Reeves, Samy, and Peterson 2015), but these approaches are sensitive to surveillance gaps, and may be prone to surprises (i.e., outbreak risk may be underestimated in areas where spillover has not been previously observed). For a more flexible approach, we examined a set of basic geospatial datasets that captured animal reservoirs and their contact patterns with humans, and aimed to identify a subset that were reflective of known outbreak history (Deka and Morshed 2018; Ramshaw et al. 2019).

Since its emergence in 1998, Nipah virus has exhibited two broad syndromes of spillover risk. Sometimes, the virus reaches humans through an intermediate host, such as pigs (in the 1998 outbreak in Malaysia) (Pulliam et al. 2012)) or horses (in the 2014 outbreak in the Philippines (Ching et al. 2015)). In Bangladesh, where Nipah virus spillover occurs more regularly, the virus has reached humans directly from bat reservoirs, usually through date palm sap contaminated with bat secretions. We chose to use the richness of known bat host species as a coarse proxy for viral circulation in reservoirs, and therefore as a reasonable first-principles approximation of risk (Plowright et al. 2019). We also explored additional predictors that captured anthropogenic drivers of spillover (McKee et al. 2021; Epstein et al. 2020; Becker et al. 2021), but found that these had poor correspondence to previous outbreaks, and excluded key areas that might be at risk (see Supporting Information).

Although MERS-like coronaviruses are found in bats worldwide (Anthony et al. 2017; Olival et al. 2020; Munoz et al. 2022), the origin of MERS-CoV appears to have been a single bat-to-camel transmission event; the virus has since jumped from camels to humans at least 200 times (Ramshaw et al. 2019), almost exclusively in livestock keeper populations. Although MERS-CoV circulates in camels throughout northern Africa, the Arabian peninsula, and even southern Asia (Dighe et al. 2019), reported spillover events have historically been confined to Saudi Arabia, Qatar, Oman, and the United Arab Emirates (Ramshaw et al. 2019). However, serological studies have found evidence of MERS-CoV exposure in Kenyan livestock handlers (Liljander et al. 2016; Kiyong’a et al. 2020), and during the preparation of the study, an active case cluster indicative of spillover was finally identified in Kenya (Ngere et al. 2022). We estimated the risk of spillover based on the average of two scaled, log-transformed predictors (Figure S2): the number of camels, based on unpublished data from the United Nations (UN) Food and Agriculture Organization (FAO)’s Gridded Livestock of the World study (Gilbert et al. 2018); and the human population employed in the agricultural sector (United Nations International Strategy for Disaster Reduction 2015).

Each predictor layer was aggregated from raster layers to an administrative polygon selected based on the spatial extent and population density in the region (state or equivalent first subnational level [i.e., ADM-1] for MERS-CoV, and second subnational administrative level [i.e., ADM-2] for Nipah). These predictor layers were rescaled between zero and one, and for MERS-CoV, the two component layers were averaged (Figure 1). Both of the final risk surfaces reasonably approximate real data on spillover risk, while identifying much broader areas where outbreaks could plausibly start someday. We therefore used these layers to drive our simulations, and seeded outbreaks based on a random multinomial sample of administrative units weighted by their scaled emergence risk.

**Figure 1.**
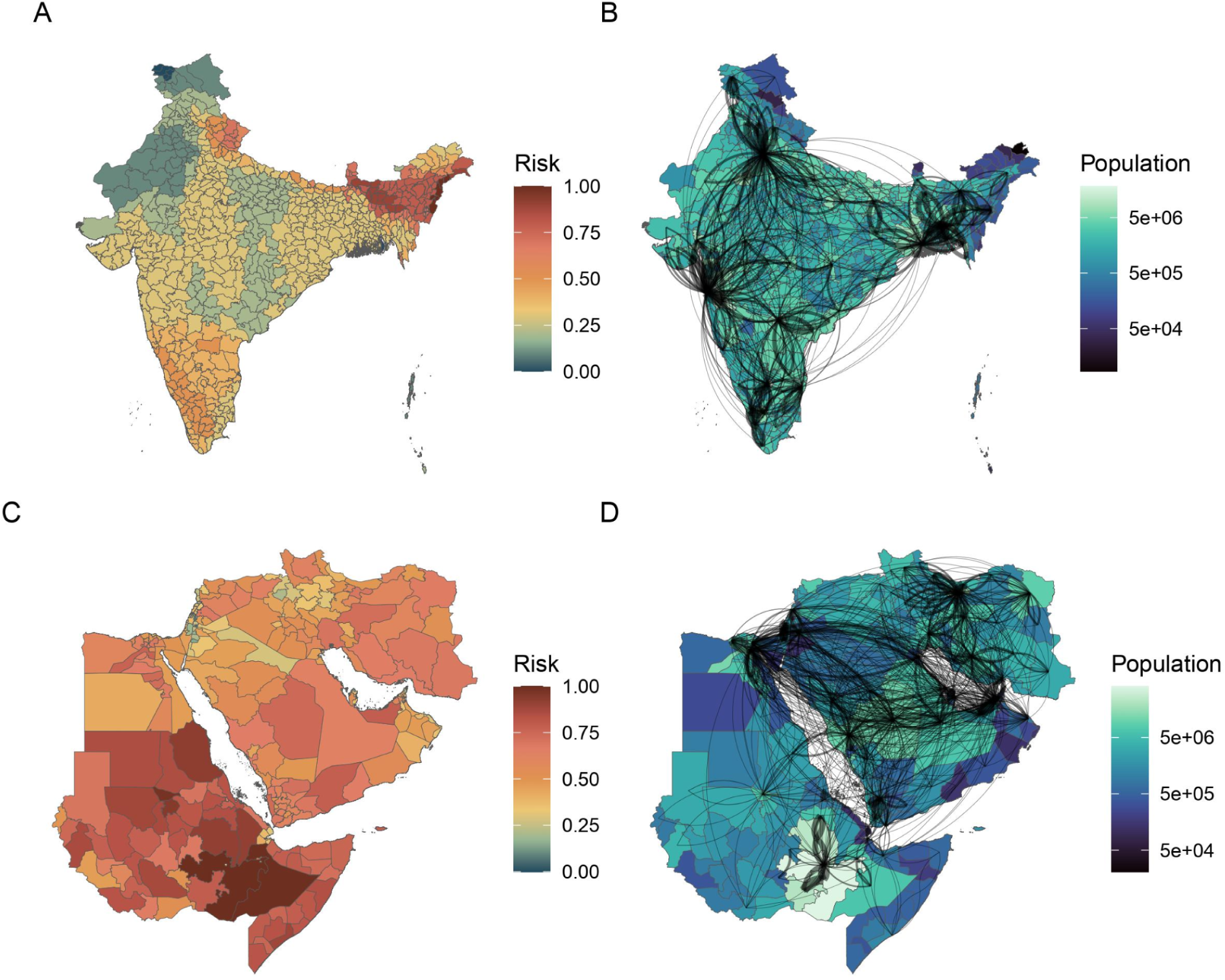
Spatial heterogeneity driving simulated epidemic dynamics. For both Nipah virus (A,B) and MERS-CoV (C,D), simulations happen in two stages: outbreaks are generated, and then spread on a human metapopulation. In the emergence stage, spillover risk is generated from mechanistic predictions and scaled from 0 to 1 (A,C). In the epidemic stage, spread is governed by population density, healthcare access, and a gravity model for traveler frequency (B,D; network edge weight indicates relative traveler volume; and only larger population centers are shown for visualization purposes). The countries included in the regions are listed in Table S1 and S2.

### Stage 2. Epidemiological dynamics post-emergence

For both pathogens, we modeled human-to-human transmission using a standard susceptible - exposed - infectious - resistant (SEIR) compartmental model, simulated on a metapopulation of administrative units (ADM-1 for MERS and ADM-2 for Nipah) with significant within-population structure. Unless otherwise specified, epidemiological parameters used in the model were estimated from clinical reports (e.g., (Assiri et al. 2013)) or gray literature (e.g., fact sheets from the U.S. Centers for Disease Control and Prevention or the World Health Organization). The same set of parameters was included in both models (Table 1), with the exception of asymptomatic rate: whereas subclinical cases of Nipah virus are generally presumed rare (Kumar et al. 2019; Nikolay et al. 2019), we assumed that a significant portion of MERS-CoV cases are likely to be subclinical (Bernard-Stoecklin et al. 2019; Grant et al. 2019; Al-Tawfiq and Gautret 2019). For these subclinical MERS cases, we assumed that they would be both non-infectious (transitioning directly from the exposed to recovered class) and functionally asymptomatic (and therefore do not trigger ring vaccination) given limited evidence for asymptomatic MERS transmission (Grant et al. 2019; Drosten et al. 2014).

**Table 1.** Key epidemiological parameter estimates used in the simulations, and parameter ranges explored in the global sensitivity analysis. (Abbreviations: HCW = healthcare worker; s.d. = standard deviation.)

| Parameter | MERS-CoV |  | Nipah virus |  |
| --- | --- | --- | --- | --- |
|  | Estimate | Range | Estimate | Range |
| Transmission rate (per day per person) ($\beta$) | 0.225 | – | 0.40 | – |
| Rate of progression to infectious (per day) | 1/6 | (1/14, 1/2) | 1/8 | (1/14, 1/5) |
| Recovery rate (per day) | 1/30 | (1/40, 1/2) | 1/10 | (1/15, 1/3) |
| Case fatality rate (per day) | 1/12 | (1/20, 1/5) | 1/3 | (1/10, 1/2) |
| Asymptomatic rate | 0.70 | (0.30, 0.90) | 0.0 | – |
| Number of contacts (mean) ($\tau$) | 11 | – | 20 | – |
| Number of contacts (s.d.) | 4 | – | 16 | – |
| Delay for contact tracing (days) | 7 | (1, 14) | 7 | (1, 14) |
| Number of missed contacts (mean) | 3 | – | 3 | – |
| Number of missed contacts (s.d.) | 5 | – | 5 | – |
| Mean time to hospital (days) | 6 | (3, 12) | 3 | (1, 10) |
| Proportion of risky contacts for HCWs | 0.05 | (0, 0.5) | 0.05 | (0, 0.5) |
| Vaccine efficacy | 0.75 | (0.30, 0.95) | 0.75 | (0.30, 0.95) |
| Time from vaccination to protection (days) | 14 | (7, 28) | 14 | (7, 28) |
| Cases to initiate vaccination (per district) | 2 | (1, 10) | 2 | (1, 10) |
| Time until vaccine availability (days) | 7 | (1, 28) | 7 | (1, 28) |
| Ring vaccination rate | 1/10 | (1/14, 1/4) | 1/10 | (1/14, 1/4) |
| Healthcare worker vaccination rate | 1/3 | (1/7, 1) | 1/3 | (1/7, 1) |

Each administrative unit was treated as a distinct population, with a total estimated size based on Gridded Population of the World version 4 (Center for International Earth Science Information Network - CIESIN - Columbia University 2016). Outbreaks spread spatially based on a gravity model (Figure 1; additional details in the Supplement), which scales probability of infection based on the distance between and population sizes of any two areas; this approach has been previously shown to be a reasonably accurate proxy for epidemic spread at large scales (Balcan et al. 2009). Outbreaks spread across populations based on either travel of infected (but not hospitalized) persons to new areas, or travel of susceptible persons from new areas to areas with ongoing outbreaks.

Within each population, we modeled multiple layers of transmission. All individuals started in the community layer (i.e., population), with a single infected case to initiate an outbreak. Frequency-dependent transmission was allowed to occur across the entire community, where new infections were generated at time *t* a rate of β*I*(*t*)/*N*. Transmission within closer contact networks was modeled as a second layer, where each new infection from community transmission would expose a new “household” (close contact network) to elevated risk, based on an average number of within-household or otherwise close contacts τ estimated from previous outbreaks of both Nipah virus and MERS-CoV (Gurley et al. 2007; Drosten et al. 2014). This higher-risk proportion of the social network expanded as a function of the cumulative number of infections from community transmission *I*(∞), and new infections were generated at a rate of β*I*(*t*)/(τ*I*(∞)).

Finally, we treated hospitalization and nosocomial transmission as a distinct layer of population structure. Infected persons were randomly hospitalized at a daily rate, based on literature estimates of average time to hospitalization for MERS-CoV in Saudi Arabia (Drosten et al. 2014) and, as a proxy for Nipah virus, malaria in India (Yadav 2010; Singh et al. 2017); new data on Nipah virus outbreaks in Bangladesh, published during our study, support this approximation (Nikolay et al. 2019). Nosocomial transmission was also assumed to be frequency dependent, where hospitalized patients *H* infected a mix of other patients and healthcare workers *M* at a rate of β*H*(*t*)/*M*. We used World Bank estimates of beds per 1,000 persons at a national level to estimate hospital capacity (World Bank 2012), and assumed 25% of beds were occupied at the start of an outbreak. We used the same source for national estimates of the number of clinicians per 1,000 persons, and assumed that supplemented with additional data from the World Health Organization and local ministries of health when necessary (e.g., for South Sudan and Palestine; Tables S1 and S2). Because no finer-scale data was available, we assumed that healthcare workers were distributed proportionally to population within countries across administrative units. We also assumed that, due to behavioral risk reduction, access to personal protective equipment, and other factors (Hadley et al. 2007), patient-clinician contact is significantly lower risk than contact within the general population.

### Stage 3. Vaccination

A number of vaccine candidates are already in various stages of development and clinical trials for both MERS-CoV and Nipah virus (Foster et al. 2022; van Doremalen et al. 2019; Bosaeed et al. 2022; Modjarrad et al. 2019; Alharbi et al. 2017; Ithinji et al. 2022), but so far, none have been licensed for use in human populations. Given the resulting uncertainty, and for modeling simplicity, we assumed a one-dose regimen for both vaccines, and that vaccination results in sterilizing immunity (vaccinated individuals are no longer susceptible), but also assume that immunity takes time to be conferred, that efficacy is less than 100%, and that there could be delays in vaccine availability after public health response is first mobilized.

In epidemic simulations, we examined three possible strategies for vaccination: (1) Vaccination of healthcare workers only: once vaccination thresholds are passed and vaccines are available, all healthcare workers are rapidly vaccinated. (2) Healthcare worker vaccination as in scenario 1, plus ring vaccination: once vaccination thresholds are passed and vaccines are available, the individuals in contact with a sick person (i.e., all those within the household layer) are vaccinated within 10 days. However, a small proportion of contacts are assumed to be missed by this process, allowing some onward transmission to slip through. (3) Healthcare worker and ring vaccination as in scenario 2, plus catch-up vaccination: after an additional delay to allow for contact tracing, all “missed contacts” are also identified and vaccinated.

### Model implementation and sensitivity analysis

Epidemic simulations were stochastic at every level, requiring a substantial number of iterations to fully sample the possible range of outbreaks. For both pathogens, we simulated 10,000 outbreaks until 10 days after the last case appeared (in communities or hospitals), or for a maximum of one calendar year (365 days), at which point simulations were interrupted. In our model, R_0_ is dependent not only on transmission rate, but also on recovery rate, death rate, and time to hospitalization. We approximated R_0_ in the community layer for a deterministic formulation of our model, and found that the R_0_ value ranged from 0.31 to 1.03 (mean ± s.d.: 0.57 ± 0.15) for Nipah virus and 0.07 to 0.41 (mean ± s.d.: 0.21 ± 0.09) for MERS-CoV. (The large difference between these estimates is due to the effect of including non-infectious asymptomatic cases of MERS-CoV.)

Because simulations had several layers of complexity, and not all parameters were equally supported by peer-reviewed literature, we conducted a global sensitivity analysis to evaluate how vaccine stockpile needs changed in response to model parameterization. We selected 12 parameters for Nipah virus (excluding asymptomatic rate, which is negligible) and 13 parameters for MERS-CoV, and chose not to examine variation in transmission rate β, given that estimates were based on data from previous outbreaks. We executed the sensitivity analysis using Latin hypercube sampling (LHS) to select 50 parameter sets with non-overlapping values from the ranges reported in Table 1. We then analyzed the results of the sensitivity analysis using generalized linear mixed models (GLMMs) to characterize how the number of vaccine doses was explained by each parameter.

## Results

### Vaccine stockpile needs

Regardless of pathogen or vaccination strategy, we found that the majority of simulated outbreaks only required a few thousand vaccine doses for containment. This was unsurprising given the epidemiology of both pathogens, which has historically been self-limiting except in rare cases. Our simulations reproduced this dynamic intentionally: in roughly a quarter of MERS-CoV simulations, and nearly a third of Nipah virus simulations, index cases failed to produce any secondary cases (and so no vaccines were deployed).

We examined the number of doses needed across a range of quantiles as a way of capturing different levels of acceptable risk for stockpile size (Table 2). Even with full ring and catch-up vaccination, only ∼2,000 doses were needed to contain a median-sized outbreak of either pathogen. However, our simulation also reproduced a strong overdispersion of outbreak sizes, where the largest epidemics were several orders of magnitude larger than the median. In the 99th percentile of outbreaks, around ∼150-180,000 doses of Nipah vaccines and ∼50,000 doses of MERS-CoV vaccines could be needed; in the maximum-sized outbreaks we simulated, up to ∼1-2 million doses of Nipah vaccines and ∼85-100,000 doses of MERS-CoV vaccines could be needed during the first year of an epidemic. These estimates were generally independent of vaccination strategy; the increased number of doses needed for more intensive strategies were generally compensated for by reductions in transmission. However, in the largest Nipah virus outbreaks, we found that community vaccination substantially reduced total needs compared to only vaccinating healthcare workers.

**Table 2.** Number of doses used at the end of one year (average across simulations).

| Percentile of outbreak size | Vaccination strategy 1:<br>healthcare workers only | Vaccination strategy 2:<br>1 + ring vaccination | Vaccination strategy 3:<br>2 + catch-up vaccination |
| --- | --- | --- | --- |
| MERS-CoV |  |  |  |
| Median | 1,236 | 1,523 | 2,089 |
| 75th | 5,944 | 6,561 | 6,826 |
| 90th | 15,191 | 15,426 | 16,321 |
| 95th | 21,096 | 20,929 | 21,647 |
| 99th | 53,638 | 48,542 | 50,212 |
| Maximum | 84,367 | 102,788 | 86,690 |
| Nipah virus |  |  |  |
| Median | 1,246 | 1,859 | 1,882 |
| 75th | 5,260 | 6,207 | 6,168 |
| 90th | 12,313 | 14,202 | 14,181 |
| 95th | 23,396 | 29,507 | 29,532 |
| 99th | 185,747 | 162,400 | 151,320 |
| Maximum | 2,105,972 | 1,019,554 | 1,128,042 |

Along this gradient of outbreak sizes, the geography of vaccination needs changed substantially (Figure 2). Larger outbreaks were more likely to reach major population centers, especially for Nipah virus, for which simulated spillover risk was independent of population density. As a result, vaccination needs in smaller outbreaks more closely tracked spillover risk, while in larger outbreaks, the greatest needs were in cities like Riyadh, Saudi Arabia or Delhi, India. In a major outbreak, vaccination needs in these cities could easily be 3-5 orders of magnitude greater than in rural areas. While this pattern reflected an obvious relationship with population density, it was also heavily driven by the geography of healthcare: for example, vaccination needs were much higher in the high-income parts of the Arabian peninsula than in comparable African cities, due to much higher numbers of healthcare workers.

**Figure 2.**
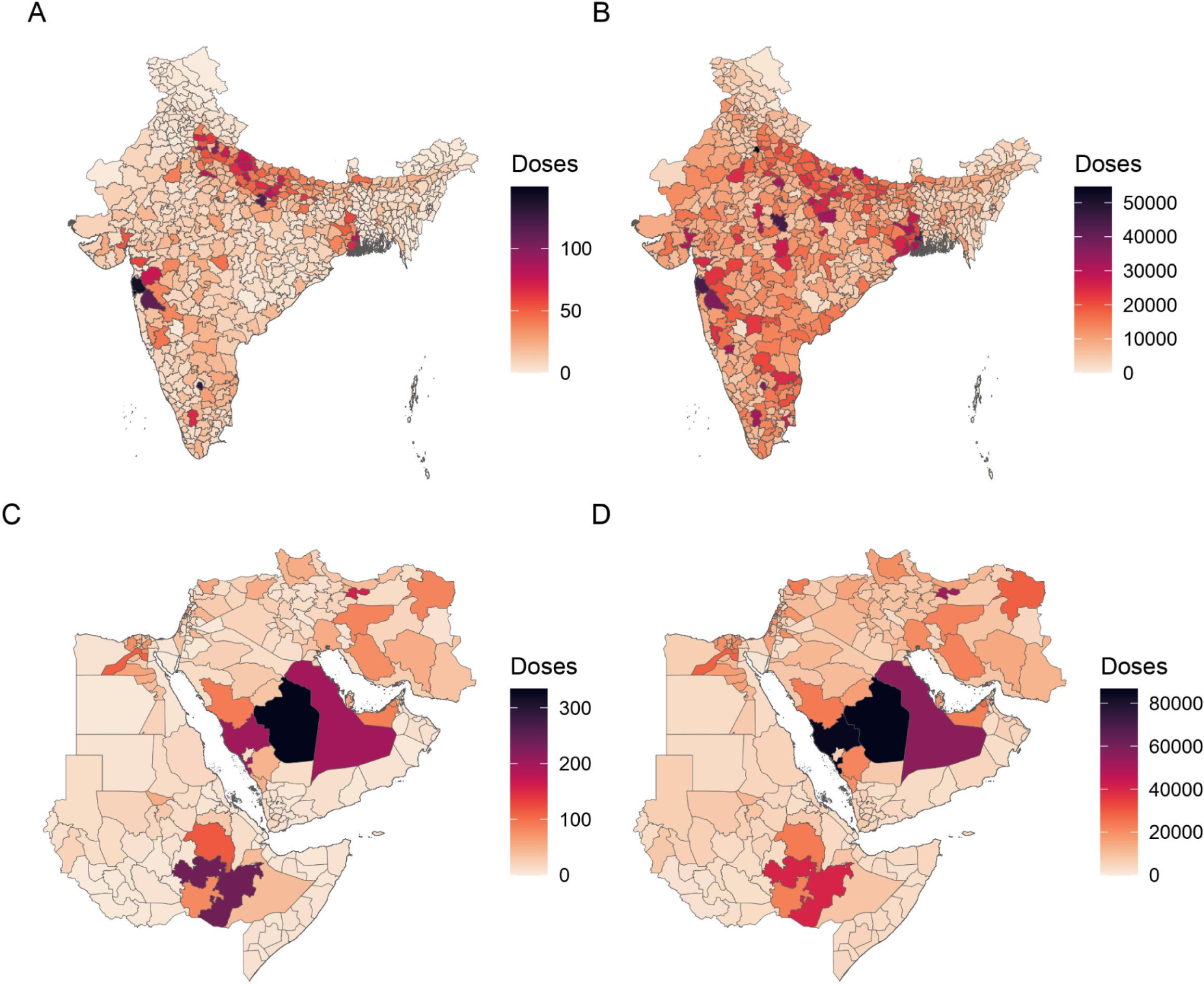
Spatial heterogeneity in predicted outcomes. Vaccination needs for Nipah virus (A,B) and MERS-CoV (C,D), based on the mean (A,C) or maximum (B,D) estimated stockpile size (including ring, catch-up, and healthcare worker vaccination) across 10,000 simulations.

### Sensitivity analysis

In both models, sensitivity analyses found that the strongest determinants of stockpile size were those that shaped R_0_ and therefore final outbreak size (Figure 3). Both a higher death rate and a higher recovery rate could substantially decrease transmission and shorten outbreak duration, while increased delays in hospitalization left more time for uninterrupted community transmission and spatial spread across administrative areas.

**Figure 3.**
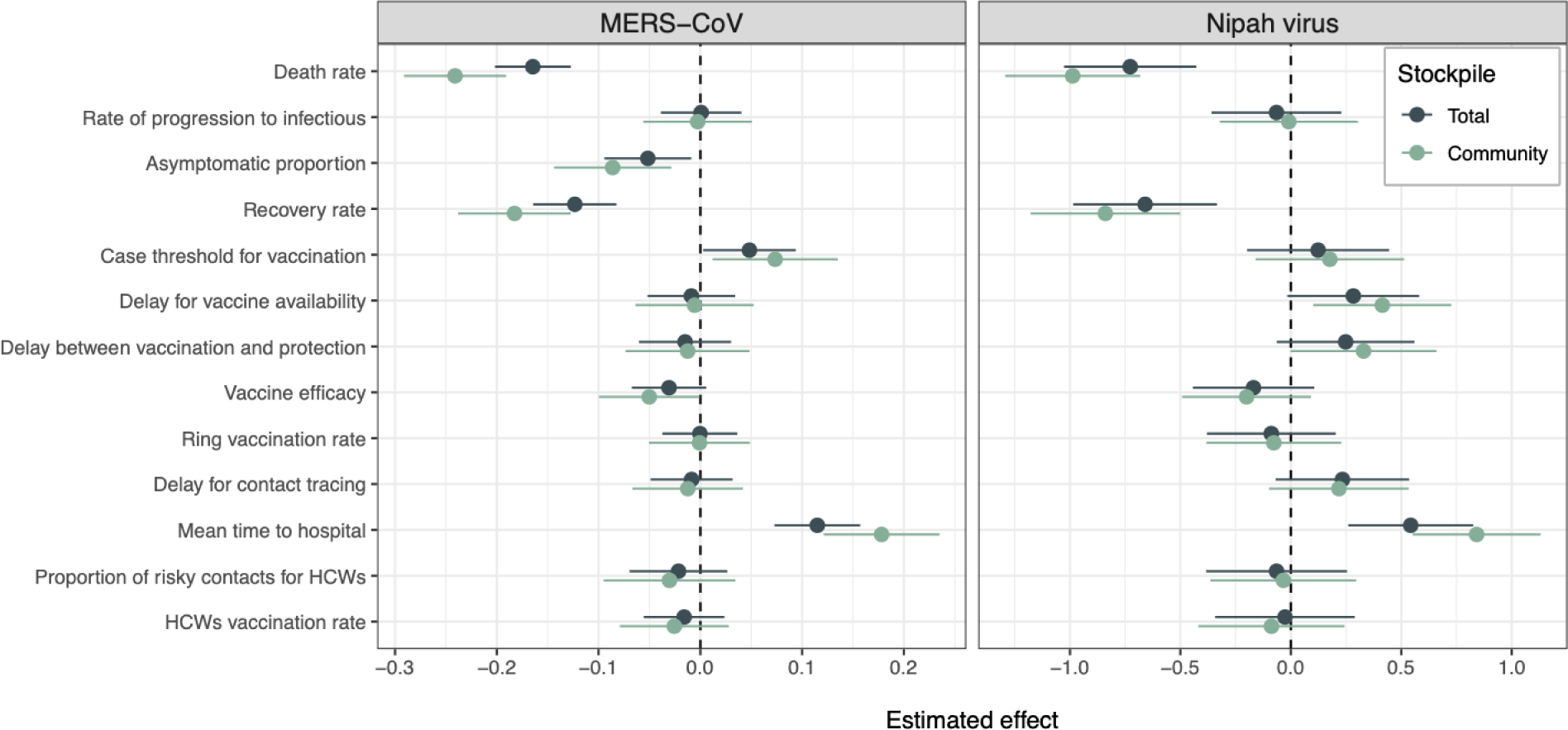
Sensitivity analysis. For each parameter, the mean estimated effect and 95% confidence interval is shown, where effects that do not overlap zero are statistically significant. Values are shown for both the full vaccine stockpile (blue) and the community stockpile (i.e., excluding healthcare worker vaccination; green).

In the Nipah virus simulations, the total community vaccination was also somewhat increased by aspects of vaccine rollout, including delays in vaccine availability and, to a minor degree, delays in protection conferred. The effect of both was even less pronounced in the full model, confirming that their effect was through vaccination mediating R_0_ in the community.

For MERS-CoV, the effects of other parameters were more idiosyncratic. Higher asymptomatic rates and, to a minor degree, higher vaccine efficacy both had a similar effect to death and recovery rates, in both cases by creating more dead ends for onward transmission. Increasing the case threshold for vaccination similarly resulted in delayed containment and increased total community vaccination needs. When healthcare worker vaccination was included, this pattern inverted, due to the design of the simulation exercise: because nosocomial transmission dominated the simulated MERS-CoV outbreaks, higher case thresholds for vaccination and longer delays to vaccine availability both reduced total doses needed, simply by shortening the period over which healthcare worker vaccination was occurring. Total vaccination needs also decreased with a higher asymptomatic rate, and to a lesser degree, with incubation period.

## Discussion

Using our multiscale modeling framework, we estimated that a few thousand doses would probably be able to suppress a typical outbreak of either pathogen, but an active stockpile of fewer than 1 million doses of MERS-CoV vaccine and at most 2-3 million doses of Nipah virus vaccine will very likely be sufficient for the first year of an acute regional epidemic. Actual needs will be determined by basic epidemiology, vaccine quality, and deployment strategy, but overall, we found that our estimates were generally robust to these uncertainties. This places our estimated needs comfortably within the range of comparable stockpiles; for example, the WHO’s global stockpiles of oral cholera vaccine and yellow fever vaccine both began with a baseline of 2 million doses, and the latter was increased to 6 million doses in 2016 to manage the growing number of outbreaks in both Africa and South America.

More broadly, our study demonstrates that stockpile design is a spatially-structured problem. A significant body of work recognizes that epidemic dynamics of many infectious diseases are heavily shaped by several aspects of human behavior, including animal-human contact (Plowright et al. 2017; Dudas et al. 2018; Gurley et al. 2017), multiscale mobility (Balcan et al. 2009; Colizza et al. 2007), social contact networks (Bansal, Grenfell, and Meyers 2007), patient-clinician interactions (Cooper, Medley, and Scott 1999; Ueno and Masuda 2008), and several facets of epidemic response. Our study establishes a parsimonious and generalizable framework for simulating these dynamics on real landscapes, parameterized with multiple sources of real-world data, without the opacity, complexity, and computational cost frequently associated with agent-based approaches (Ajelli et al. 2010; Tracy, Cerdá, and Keyes 2018). While our quantitative predictions are dependent on the model structure and parameters driven by data availability, our work qualitatively highlights that a spatially-structured emergence and transmission risk landscape leads to geographically heterogeneous stockpile needs.

Developing this modeling framework also highlights how scientific uncertainty is unevenly distributed across different stages of the disease emergence process (Pigott et al. 2017). For both pathogens, we found that mechanistic proxies had a limited ability to generate realistic landscapes of spillover risk. Datasets cataloging prior spillovers are also workable as a proxy for risk (Lerch et al. 2022), but would have failed to anticipate major recent outbreaks of both pathogens we examined here (Arunkumar et al. 2019; Ngere et al. 2022). In the future, global databases of wildlife disease prevalence could be used to develop more precise maps of spillover risk. However, our simulations suggested that this layer of uncertainty is not the most important for stockpile design; in a regional emergency, optimal vaccine allocation will mostly need to track population density and healthcare availability. This finding tracks with previous work simulating Ebola virus epidemics, which found that only 14% of variation in final spatial extent was determined by outbreak origin (Kramer et al. 2016).

Within the process of human-to-human transmission, we found that the strongest predictors of final epidemic size (and therefore vaccine stockpile needs) were basic disease characteristics like case fatality and symptomatic rates. This finding underscores the importance of descriptive epidemiology (Lesko, Fox, and Edwards 2022; Fox et al. 2022), and the value of studies that revisit historical outbreaks, in order to better calibrate stockpile needs. Ability to access healthcare was also a key determinant of outbreak progression, underscoring the risk that devastating epidemics will start at the edge of surveillance systems and go undetected for weeks to months (Glennon et al. 2019; Weiss et al. 2020; Hulland et al. 2019). In contrast, we found that stockpile needs were largely insensitive to the specific features of different vaccines or their distribution. While basic principles of outbreak response generally hold—vaccines that confer sterilizing immunity at a high rate are the most useful for containing outbreaks, and effective community vaccination will best reduce the burden on healthcare systems—a sufficiently large stockpile of any vaccine could be an essential life-saving countermeasure in the coming decade. Given concerns that more transmissible viral variants or relatives exist in reservoirs, and the high probability of another pandemic comparable to Covid-19 in the coming decades (Marani et al. 2021), the ethical imperatives and financial incentives to establish these stockpiles are substantial. With future progress on the global health security challenges of vaccine equity, vaccine hesitancy, supply chain and cold chain infrastructure, and global health data sharing and coordination, vaccine stockpiles can hold the promise of rapid and effective response to emerging disease outbreaks.

## Data Availability

No original data are used in the study.

## Acknowledgements

This project was made possible with financial support provided by the Coalition for Epidemic Preparedness Innovations. We thank Alison Bettis, Georgina Murphy, and Mauro Bernuzzi of CEPI for their support during the development of our modeling framework. We are grateful to Emily Gurley (Nipah) and Bart Haagmans (MERS) for serving as pathogen experts, to Steph Seifert and Dan Becker for thoughtful feedback on the project, and to Tim Robinson and Marius Gilbert for sharing unpublished data on camel population density. CJC was additionally supported by NSF BII 2213854, and thanks colleagues in the Verena program for thoughtful conversations about mapping landscapes of zoonotic risk.

## Author Contributions

SB, RG, and CJC designed the study. RG, CJC, and AT collected necessary data. RG, CJC, AT and SB ran all analyses and interpreted the findings. CJC, SB, and RG wrote the manuscript, and all authors contributed to editing and approved the manuscript.

## Data and Code Availability

Code to reproduce the analyses is available at github.com/bansallab/vaccinestockpile.

## SUPPORTING INFORMATION

### Text S1. Appendix

#### I. Additional details on the mechanistic emergence risk maps

##### Alternate constructions of the Nipah virus risk layer

In Bangladesh, the emergence of Nipah virus has been driven by the conversion of intact forest into mosaic landscapes, driving the flying fox bat (*Pteropus medius*) reservoir into farms, where they forage for fruit and date palm sap (McKee et al. 2021; Epstein et al. 2020). These foraging behaviors create opportunities for human exposure to Nipah virus, and probably coincide with seasonal patterns of winter temperature, food stress, and site selection—a pattern that has been demonstrated more comprehensively for *Pteropus* bat reservoirs of Hendra virus in Australia (Becker et al. 2021; Eby et al. 2023; McKee et al. 2021; Epstein et al. 2020).

In an effort to capture these patterns, we originally considered six possible components for the mechanistic Nipah virus emergence risk layer (Figure S1):

- species richness of known bat hosts (Plowright et al. 2019);
- a proxy for bat “abundance,” or at least the frequency of human-bat interactions, derived from the density of records for the order Chiroptera in the Global Biodiversity Informatics Facility online database (www.gbif.org);
- tree cover in the year 2000 (Hansen et al. 2013) and tree cover loss by 2019 (accessed from the Global Forest Watch data portal: globalforestwatch.org);
- palm fruit production (by volume) and palm oil production (by cultivated area), approximated based on source datasets by country: cultivated area up to March 2017, and provisional fruit production between 2016 and 2017, by ADM1 region in India (Directorate of Economics & Statistics 2018); and area under garden from 2016 to 2017, and total fruit production from 2016 to 2017, by ADM2 region in Bangladesh (Bangladesh Bureau of Statistics 2018).

None of these layers was particularly concordant with historical outbreak geography; most suggested that spillover rates should be high in north eastern India and Andhra Pradesh, neither of which have had a recorded outbreak. More significantly, other than fruit production, most of these layers failed to capture the high rate of spillover in Bangladesh and northern West Bengal. Given those limitations, we chose to only use bat host species richness as a first principles proxy for viral presence or absence in wildlife (Plowright et al. 2019).

##### Technical validation

Although our predictions are mechanistic and independent of spillover data, we examined correspondence with real spillover datasets as a technical validation step. For Nipah virus, all recorded spillover events in south Asia have occurred in Bangladesh or West Bengal, with the notable exceptions of the 2018 and 2023 outbreaks in Kerala (Pigott, *pers. comm.*). This is reasonably well-captured by the bat richness model, although areas of high host richness in northern and north eastern India are notably devoid of known outbreaks. Sampling in both of these regions has been limited, but seropositive bats have been recorded in Haryana (Epstein et al. 2008), suggesting that spillover risk in northern India could be nontrivial; north eastern India is similarly situated adjacent to the spillover hotspot in Bangladesh and well within the known extent of viral circulation in south and southeast Asia (Plowright et al. 2019).

For MERS-CoV, our risk surface captures the spillover hotspot in the central Arabian peninsula reasonably well (Ramshaw et al. 2019). Our model does imply a reasonably high risk of spillover in Sudan, Ethiopia, Somalia, Iran, Syria, and parts of Egypt. Given the high seropositivity of camels across the region (Dighe et al. 2019), and recent evidence of zoonotic transmission in Kenya (Ngere et al. 2022; Kiyong’a et al. 2020), spillover risk should be considered plausible throughout the entire region captured in our emergence risk layer.

#### II. Additional details on the gravity model

To capture the probability of infection driven by human mobility within countries for both diseases, we adapted a model used by Kramer *et al*. (Kramer et al. 2016), who found the model to be the best fit for spatial infection dynamics during the West Africa Ebola outbreak. Here the probability of infection between a susceptible location *i* and source location *j* is a logistic function of the Euclidean distance between centroids of location *i* and *j* (*d_ij_*), and is weighted by the population sizes of the two locations:

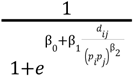

We parameterized the model as follows based on Kramer *et al*.’s results:

β_0_ = 5. 166, β_1_ = 157. 1, β_2_ = 0. 189. Additionally, we limited cross-country movements to administrative units that shared a border, and added a penalty to the gravity-model driven probability of infection for cross-country movements. These rulesets were necessary to appropriately capture the dynamics of a “regional epidemic” spreading mostly through ground travel (similar to, e.g., the 2014 outbreak of Ebola in West Africa). Although air travel could enable broader international spread—as in the case of the 2015 outbreak of MERS-CoV in South Korea—a full pandemic simulation was beyond the scope of the present work; airports and international travelers are also increasingly well-surveilled for emerging pathogens (Ahmed et al. 2022; La Rosa et al. 2022).

#### III. Additional details on the sensitivity analysis

In the sensitivity analysis, for each of the 50 parameter sets, we performed 1,500 stochastic simulations, or a total of 75,000 additional outbreak simulations, using the full vaccination regime (strategy 3). Because our aim was to understand inter-outbreak variability in a controlled environment, we only ran simulations for 180 days or until 10 days after the last case, and restricted outbreak seeds to the top 10 highest-risk areas for emergence; for MERS-CoV, to emulate previous outbreaks (Ramshaw et al. 2019), we further constrained these locations to the Arabian peninsula.

We analyzed these simulation results using generalized linear mixed models (GLMMs), with the number of vaccine doses as the response variable (modeled using a negative binomial link function, given overdispersion of outbreak sizes) and the different parameter values used as explanatory predictors. Because we were only interested in the variability of stockpile needs, we restricted the analysis to simulations where outbreaks spread past the initial introduction and triggered vaccination; we also analyzed doses with and without including healthcare worker vaccination. Models included a random effect of the parameter set (each of the 50 combinations), and another for the emergence location, to account for different population sizes in different locations resulting in different vaccine usage.

**Figure S1.**
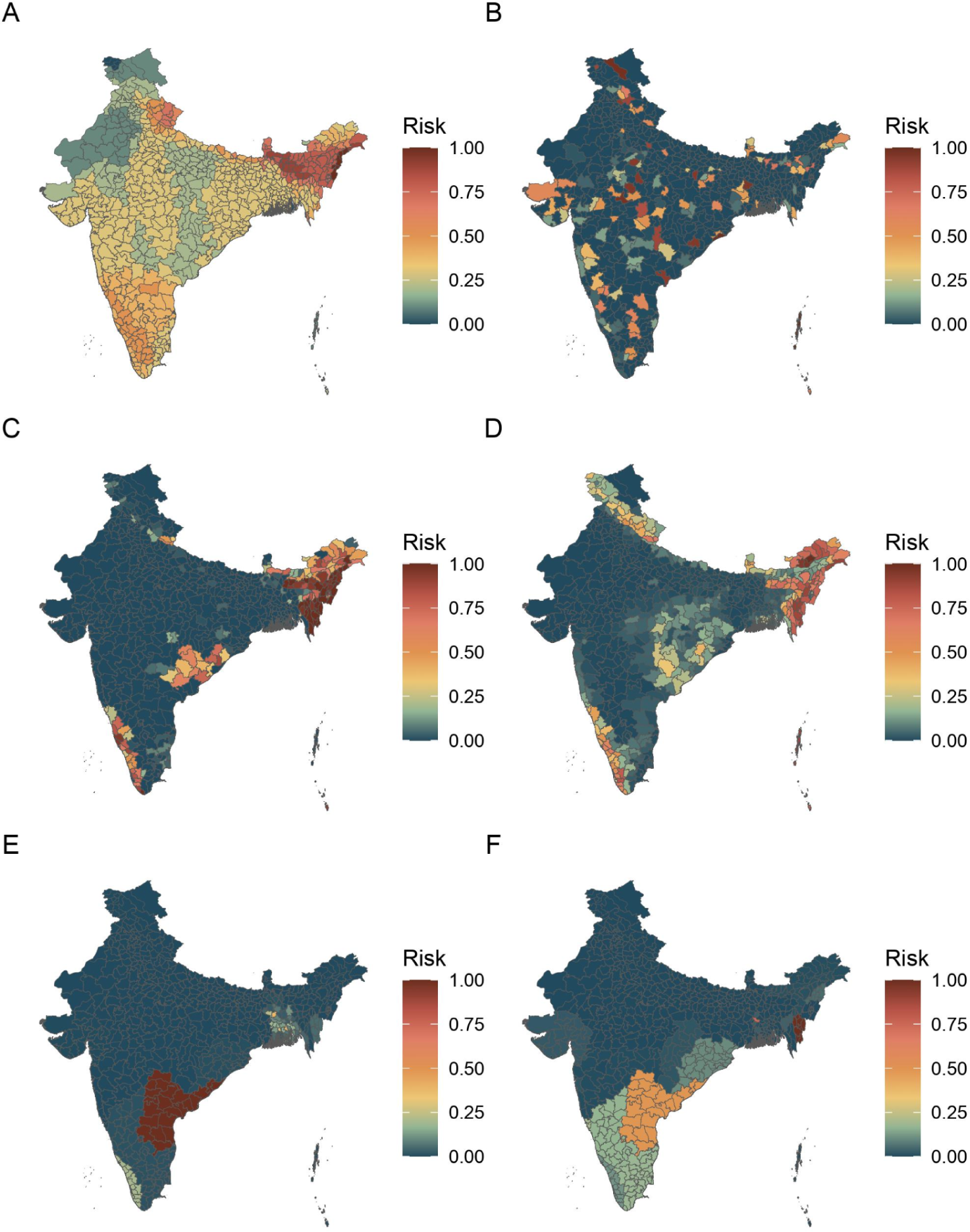
Components considered for the Nipah virus emergence risk layer. All layers are rescaled between 0 and 1. (A) Species richness of known bat hosts of Nipah virus. (B) Density of bat occurrences reported in a biodiversity database (as a proxy for both bat populations and bat-human contact). (C) Deforestation rates. (D) Tree cover. (E) Palm agriculture production for fruit. (F) Palm agriculture production for palm oil.

**Figure S2.**
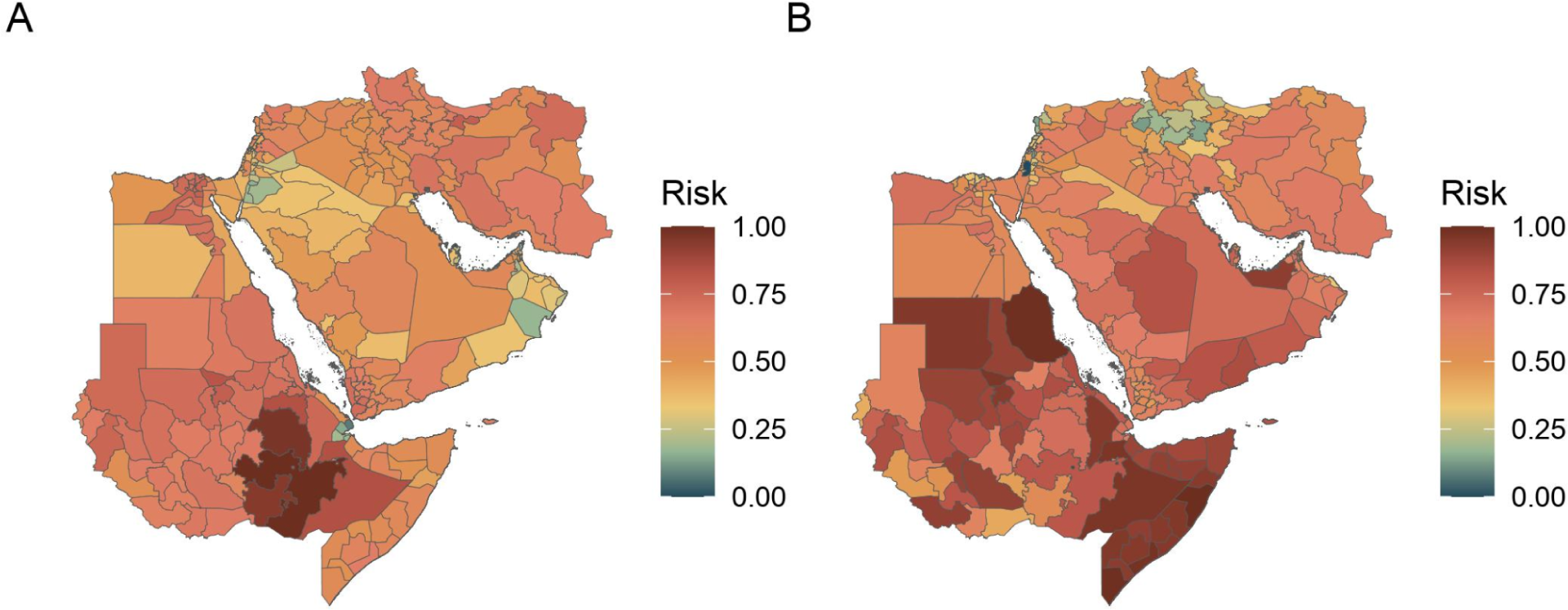
Components of the MERS-CoV risk surface. Both layers are log-transformed and rescaled between 0 and 1. (A) Human population working in the agricultural sector. (B) Estimated camel populations. The two layers are averaged in the final emergence layer.

**Table S1.**
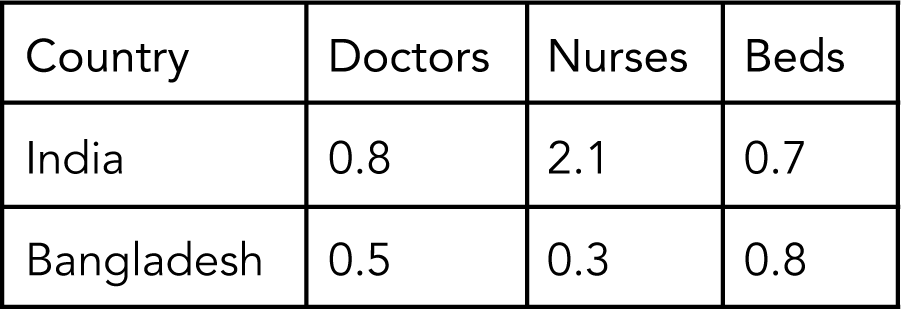
Number of doctors, nurses, and beds per 1,000 inhabitants by country in the Nipah virus model. Data from the World Bank.

**Table S2.** Number of doctors, nurses, and beds per 1,000 inhabitants by country in the MERS-CoV virus model. Data from the World Bank, the WHO, and national ministries of health.

| Country | Doctors | Nurses | Beds |
| --- | --- | --- | --- |
| Bahrain | 0.9 | 2.5 | 2 |
| Djibouti | 0.2 | 0.5 | 1.4 |
| Egypt | 0.8 | 1.4 | 1.6 |
| Eritrea | 0.1 | 0.6 | 0.7 |
| Ethiopia | 0.1 | 0.8 | 0.3 |
| Iran | 1.1 | 1.9 | 1.5 |
| Iraq | 0.8 | 1.7 | 1.4 |
| Israel | 3.2 | 5.2 | 3.1 |
| Jordan | 2.3 | 3.4 | 1.4 |
| Kuwait | 2.6 | 7 | 2 |
| Lebanon | 2.3 | 2.6 | 2.9 |
| Oman | 2 | 4.3 | 1.6 |
| Palestine | 1 | 1.4 | 1.3 |
| Qatar | 7.7 | 6.6 | 1.2 |
| Saudi Arabia | 2.4 | 5.7 | 2.7 |
| Somalia | 0 | 0.1 | 0.9 |
| South Sudan | 0.01 | 0.01 | 0.1 |
| Sudan | 0.4 | 0.8 | 0.8 |
| Syria | 1.2 | 1.5 | 1.5 |
| United Arab Emirates | 2.4 | 5.6 | 1.2 |
| Yemen | 0.3 | 0.7 | 0.7 |

